# Effectiveness of COVID-19 vaccination and prior infections to reduce long COVID risk during the pre-Omicron and Omicron periods

**DOI:** 10.1101/2025.07.02.25330616

**Authors:** Sara Carazo, Jonathan Phimmasone, Danuta M Skowronski, Katia Giguère, Manale Ouakki, Denis Talbot, Charles-Antoine Guay, Chantal Sauvageau, Nicholas Brousseau, Gaston De Serres

## Abstract

**Background:** We estimated vaccine effectiveness (VE) against COVID-19 and long COVID during pre-Omicron and Omicron periods, by number of doses and prior infection history.

**Methods:** We combined survey information from a cohort of healthcare workers in Quebec, Canada, with immunization registry and laboratory administrative data. We defined COVID-19 cases as symptomatic laboratory-confirmed infections and long COVID as self-reported symptoms persisting ≥12 weeks. We assessed VE against COVID-19 and long COVID, stratified by infection history, using a test-negative design where vaccinated participants were compared to unvaccinated participants during the pre-Omicron period or to those twice vaccinated ≥6 months before laboratory testing during the Omicron period.

**Results:** Analyses included 8230 COVID-19 participants and 43361 tested specimens. During the pre-Omicron period, one- and two-dose VE was 75% (95%CI:64-83) and 95% (95%CI:84-98) against COVID-19, respectively, and 91% (95%CI:79-96) and 87% (95%CI:22-98) against long COVID, respectively. During the Omicron period, booster dose VE was 41% (95%CI:34-47) against COVID-19 and 57% (95%CI:46-66) against long COVID, waning by 6 months. Hybrid VE in vaccinated and previously infected individuals ranged 81% (95%CI:38–94) to 92% (95%CI:87–95) regardless of number of doses, prior infecting variant or median time since last immunological event up to 9 months.

**Conclusion:** COVID-19 vaccination prevented long COVID during the pre-Omicron period and reduced the risk by more than half post-Omicron. With most of the population by now both vaccinated and infected, repeated booster doses may add little incremental value against long COVID, an observation with important public health, immunization program and cost implications.

## INTRODUCTION

Long COVID is a chronic post-infectious condition that presents as an ongoing, relapsing or progressive condition that generally impacts daily functioning [1]. It is estimated that over 400 million people worldwide suffer from long COVID [2].

Most studies evaluating the protective effect of vaccination on long COVID risk assessed it exclusively among COVID-19 cases, ignoring earlier (upstream) risk reduction through the prevention of SARS-CoV-2 infection, including severe COVID-19, in the first place [3,4]. Since the emergence of Omicron, vaccine effectiveness (VE) against infections appears lower and shorter lasting [5]. While the few studies for which long COVID was considered a severe COVID-19 outcome found low to moderate protection against long COVID, data on the effectiveness of hybrid (vaccine-plus-infection induced) immunity are scarce [6–8].

In Canada, as in other jurisdictions, COVID-19 vaccination program’s goal has been to prevent severe illness and, due to short-lived vaccine protection, booster doses have been administered every 6-12 months [9,10]. In the current context of ongoing transmission of new Omicron subvariants and the high frequency of reinfections, it remains essential to measure ongoing booster dose protection against long COVID.

In spring 2023, we conducted an online survey within a cohort of Quebec healthcare workers (HCW) to evaluate long COVID frequency, risk and protective factors [11]. Combining survey data with administrative registries, we estimated VE against COVID-19 and against long COVID by number of doses (one to five), prior infection history and circulating SARS-CoV-2 variants. For comparability with other studies, we also measured vaccine-associated risk reduction among COVID-19 cases.

## METHODS

### Study design

We evaluated VE against COVID-19 using the test-negative design (TND) and estimated vaccine-associated reduction in long COVID risk using a retrospective cohort design (RCD).

### Population

Adult HCWs residing in the province of Quebec, Canada, were invited to participate in an on-line survey if working in the health and social services public network and/or registered with a professional order for physicians, nurses, nursing assistants, respiratory therapists, pharmacists, or midwives during the pandemic and able to communicate in French or English. The study population consisted of survey participants who were tested for SARS-CoV-2 by nucleic acid amplification test (NAAT) between January 3, 2021 (three weeks after vaccine campaign launch in Quebec) and February 20, 2023 (allowing 12-week follow-up for long COVID case ascertainment). We excluded participants with COVID-19: if their first self-reported COVID-19 was not confirmed by a positive NAAT in the laboratory registry (allowing a lag of 60 days between self-reported and laboratory date); if survey responses did not allow long COVID identification or attribution to a COVID-19 episode; (c) if vaccinated with >5 doses, or if last dose was not an mRNA vaccine, or if <3 months between doses or between vaccination and sampling. For hybrid immunity analyses, participants were also excluded if their second self-reported COVID-19 infection was not confirmed by a positive NAAT in the laboratory registry or if they reported long COVID after their first infection.

### Procedures

Between May 16 and June 15, 2023, participants self-reported sociodemographic and clinical data. Questions included the number, date, laboratory confirmation and severity of COVID-19 episodes, as well as persistence of at least one symptom beyond 12 weeks post-COVID-19 infection [11]. Using a unique identifier, survey data were linked to provincial health administrative databases, including: the immunization registry; the Quebec integrated chronic disease surveillance system; and the laboratory registry containing all SARS-CoV-2 NAAT performed since pandemic onset. In Quebec, HCWs were extensively tested by NAAT the first year of the pandemic and they continued to have easy access to NAAT during the Omicron period.

Variant dominant periods pre-Omicron (from January 3, 2021 to December 25, 2021) and Omicron (from December 26, 2021 to February 20, 2023) were assigned based on Quebec viral genetic surveillance as described elsewhere [12]. During the study period, the main circulating Omicron subvariants were BA.1, BA.2, BA.4/5, BQ.1 and XBB.1.5 (Figure 1).

**Figure 1.**
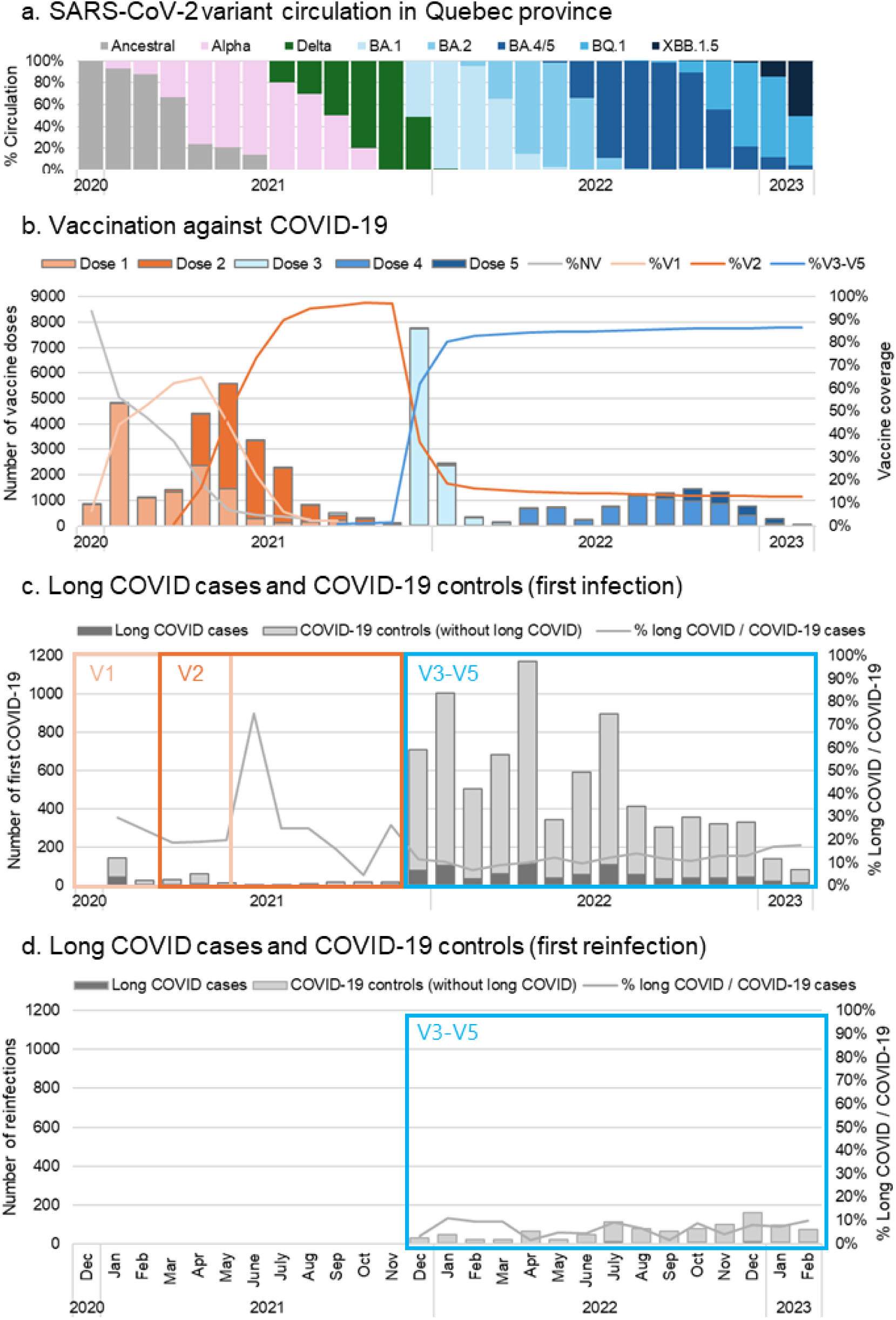
Temporal distribution of SARS-CoV-2 variant circulation, COVID-19 vaccination, long COVID cases and first COVID-19 infections and reinfections. Abbreviations: NV, not vaccinated; V1-V5, vaccination with one to five doses.

### Exposure and outcome definitions

Vaccination status was defined relative to the date of specimen collection. Participants (or tests) were considered vaccinated with one to five doses (V1 to V5) if they received a dose of mRNA vaccine (BNT-162b2 (Pfizer-BioNTech) or mRNA-1273 (Moderna)) ≥14 days (V1) or ≥7 days (V2-V5) before the NAAT date, and not-vaccinated (NV) if no COVID-19 vaccination was administered. Hybrid immunity was defined by the number of vaccine doses received (V2-V5) and a prior NAAT-confirmed pre-Omicron or Omicron infection, without long COVID.

COVID-19 cases were positive on NAAT performed for COVID-19 symptoms. First reinfection was defined as a positive NAAT performed ≥90 days after first positive NAAT. Long COVID cases were individuals self-reporting persistence of post-COVID-19 symptom(s) (among a list of 21) attributed to COVID-19 (first infection or first reinfection) that was identified as NAAT-confirmed in the laboratory registry. Moderate or severe long COVID was defined by the presence of ≥1 persistent moderate or severe symptom.

For TND estimation of VE against COVID-19 and long COVID, eligible test-negative controls were symptomatic NAAT-negative participants before a first (if any) positive NAAT. Negative tests performed ≥90 days post-first infection before the first (if any) reinfection were also eligible for the hybrid protection’s evaluation. For RCD estimation of vaccine-associated risk reduction among COVID-19 cases, controls were individuals with a SARS-CoV-2 infection (first infection or first reinfection) without long COVID (COVID-19 controls). A summary of methods is presented in Supplementary Figure 1.

### Statistical analyses

Based on vaccine doses administered and variant evolution, three distinct periods were analyzed (Figure 1) [13,14]. We evaluated one-dose vaccination (vs. NV) using specimens tested between January 3 (epi-week W2021-01) and June 12, 2021 (W2021-23) when ancestral strain and Alpha variants circulated. We evaluated two-dose vaccination (vs. NV) using specimens tested between March 28 (W2021-13) and December 11, 2021 (W2021-49), when the ancestral strain, Alpha and Delta variants circulated. We evaluated booster doses (V3-V5) and hybrid immunity (vs. V2 ≥6 months before test date, without documented or self-reported prior infection) using specimens tested between December 19, 2021 (W2021-51) and February 20, 2023 (W2023-08), when Omicron circulated. The V2 comparison group was chosen for booster dose evaluation given paucity of unvaccinated participants by the Omicron period and waned protection against Omicron by six months post-vaccination [15]. Successive booster dose protection was assessed by comparing each dose with the previous dose administered ≥6 months before the test date. These sub-analyses included tests conducted since W2021-51 (V3), W2022-14 (V4), and W2022-35 (V5). We further stratified by time since last booster vaccination in two-month intervals.

For TND VE estimation, we used incidence density sampling to randomly select ten test-negative controls (V1 and V2 analyses) or up to five test-negative controls (V3-V5 analysis) per case for each two-week period. Odds ratio (OR) and 95% confidence intervals (95%CI) were estimated by conditional logistic regression comparing test-positive COVID-19 cases and long COVID to test-negative controls, adjusting for sex, age group, region of residence, occupation, workplace, material and social deprivation indexes, and number of pre-existing comorbidities. For RCD estimation of long COVID risk reduction, OR and 95%CI were estimated by logistic regression models comparing long COVID cases and COVID-19 controls (i.e., COVID-19 cases without long COVID) adjusting for the same above-mentioned variables and for four-week periods. Absolute (V1 and V2 vs NV) and relative (V3-V5 vs V2) VE and risk reduction were calculated as: (1-OR_adjusted_)*100.

### Ethical considerations

This study was conducted under the legal mandate of the National Director of Public Health of Quebec under the Public Health Act and was approved by the Research Ethics Board of the Centre hospitalier universitaire de Québec-Université Laval.

## RESULTS

Among 400,222 Quebec HCWs invited, 22,496 (5.6%) responded to the survey. Among them, 10,281 (45.7%) SARS-CoV-2 infected individuals were linked to a corresponding NAAT in the laboratory registry, 7679 (34.1%) were not linked, and 4536 (20.2%) were uninfected individuals (Supplementary Figure S2). After exclusions, 43,361 and 38,653 NAAT were, respectively, eligible for COVID-19 and long COVID VE analyses by TND, while 8230 individuals with a first infection and 923 with a first reinfection were included in the RCD risk reduction analyses.

Case and control characteristics were globally similar, with most participants being white and middle-aged women (Table 1). More long COVID cases than COVID-19 controls had ≥2 comorbidities (217 [22.7%] vs. 1217 [16.7%]).

**Table 1.** Characteristics of cases and controls.

|  | Long COVID cases | Test-positive COVID-19 cases | Test-negative controls | COVID-19 controls (without long COVID) |
| --- | --- | --- | --- | --- |
| N | 954 | 5662 | 37,699 | 7276 |
| Characteristics | N (%) | N (%) | N (%) | N (%) |
| Sex |  |  |  |  |
| Female | 834 (87.4) | 4846 (85.6) | 32563 (86.4) | 6161 (84.7) |
| Male | 120 (12.6) | 816 (14.4) | 5136 (13.6) | 1115 (15.3) |
| Age, years |  |  |  |  |
| Median (IQR) | 43 (36 – 52) | 42 (34 – 50) | 41 (33 – 49) | 45 (38 – 53) |
| 18-39 | 288 (30.2) | 2370 (41.9) | 17481 (46.4) | 2829 (38.9) |
| 40-49 | 332 (34.8) | 1765 (31.2) | 10830 (28.7) | 2317 (31.8) |
| 50-59 | 240 (25.2) | 1146 (20.2) | 7142 (18.9) | 1522 (20.9) |
| ≥60 | 94 (9.9) | 381 (6.7) | 2246 (6.0) | 608 (8.4) |
| Race / Ethnicity |  |  |  |  |
| White | 812 (85.1) | 4994 (88.2) | 33606 (89.1) | 6448 (88.6) |
| Black | 41 (4.3) | 193 (3.4) | 1461 (3.9) | 239 (3.3) |
| Asiatic | 24 (2.5) | 142 (2.5) | 666 (1.8) | 173 (2.4) |
| Hispanic | 22 (2.3) | 93 (1.6) | 542 (1.4) | 124 (1.7) |
| Other / Not reported | 55 (5.8) | 240 (4.2) | 1424 (3.8) | 292 (4.0) |
| Comorbidity <sup>a</sup> |  |  |  |  |
| None | 491 (51.5) | 3292 (58.1) | 21142 (56.1) | 4371 (60.1) |
| One | 246 (25.8) | 1385 (24.5) | 9474 (25.1) | 1688 (23.2) |
| Two or more | 217 (22.7) | 985 (17.4) | 7083 (18.8) | 1217 (16.7) |
| Profession |  |  |  |  |
| Nursing staff, physicians, other health professionals | 536 (56.2) | 3552 (62.7) | 25391 (67.4) | 4648 (63.9) |
| Health-assisting occupations | 239 (25.1) | 1071 (18.9) | 7967 (21.1) | 1289 (17.7) |
| Management and administrative staff | 179 (18.8) | 1039 (18.4) | 4341 (11.5) | 1339 (18.4) |
| Workplace |  |  |  |  |
| Hospital | 437 (45.8) | 2753 (48.6) | 21496 (57.0) | 3511 (48.3) |
| Long-term care facilities | 232 (24.3) | 649 (11.5) | 6248 (16.6) | 1786 (24.5) |
| Clinics | 146 (15.3) | 861 (15.2) | 3804 (10.1) | 768 (10.6) |
| Other <sup>b</sup> | 139 (14.6) | 1400 (24.7) | 6151 (16.3) | 1211 (16.6) |
| Variant of first infection |  |  |  |  |
| Pre-Omicron | 105 (11.0) | 356 (6.3) | 17285 (45.9) | 337 (4.6) |
| Omicron | 849 (89.0) | 5306 (93.7) | 20414 (54.1) | 6939 (95.4) |
| Vaccination status |  |  |  |  |
| Not vaccinated (NV) | 68 (7.1) | 213 (3.8) | 4147 (11.0) | 208 (2.9) |
| Vaccinated with 1 dose (V1) | 10 (1.0) | 55 (1.0) | 5956 (15.8) | 74 (1.0) |
| Vaccinated with 2 doses (V2) | 224 (23.5) | 1175 (20.8) | 10170 (27.0) | 1247 (17.1) |
| Vaccinated with 3 doses (V3) | 527 (55.2) | 3526 (72.8) | 15768 (48.4) | 4743 (65.2) |
| Vaccinated with 4 doses (V4) | 105 (11.0) | 593 (12.2) | 1441 (4.4) | 872 (12.0) |
| Vaccinated with 5 doses (V5) | 20 (2.1) | 100 (2.1) | 217 (0.7) | 132 (1.8) |
| Type of last booster dose |  |  |  |  |
| Monovalent (V3-V5) | 615 (64.5) | 16981 (52.1) | 4022 (83.0) | 5473 (75.2) |
| Bivalent (V4-V5) | 37 (3.9) | 445 (1.4) | 197 (4.1) | 274 (3.8) |

<sup>a</sup> Included comorbidities: depressive disorder, alcohol and drug abuse, hypertension, chronic lung disease, hypothyroidism, kidney disease, fluid and electrolyte disorders, peripheral vascular disorders, obesity, dementia, cerebrovascular disease, neurological disorders, liver disease, psychosis, pulmonary circulation disorders, rheumatoid arthritis, coagulopathy, weight loss, paralysis, ulcerative disease, AIDS/HIV, anemia, cardiovascular disease, diabetes, immune system problems, cancer.
<sup>b</sup> Rehabilitation centers, patients' homes, telework, other
Abbreviations: IQR, interquartile range; NV, not vaccinated; V1-V5, vaccinated with 1 to 5 doses.

Vaccination status varied with time (Figure 1). In December 2021, 96.9% of participants were two-dose vaccinated and <1% were unvaccinated. Booster vaccine coverage achieved 82.2% by February 2022 with 13.7% of HCWs not receiving any booster dose during the study period. Overall, most first infections were due to Omicron: 849 (89.0%) among long COVID cases, 5306 (93.7%) among test-positive COVID-19 cases and 6939 (95.4%) among COVID-19 controls (Table 1). Conversely, 17,285 (45.9%) eligible negative tests were from the pre-Omicron period. During the Omicron period, 18 (2.1%) and 43 (5.1%) long COVID cases had, respectively, pre-Omicron and Omicron prior NAAT-confirmed infections, being 267 (3.6%) and 589 (8.0%) for COVID-19 controls (Supplementary Table 1).

Absolute one-dose VE estimated by TND was 75% (95%CI 64–83) against COVID-19, increasing to 91% (95%CI 79–96) against long COVID, with RCD-estimated reduction in the long COVID risk among vaccinated, COVID-19 infected HCWs of 77% (95%CI 38–82) (Table 2). Two-dose VE by TND was 95% (95%CI 84–98) against COVID-19 and 87% (95%CI 27–98) against long COVID, with no RCD evidence of long COVID risk reduction among infected HCWs.

**Table 2.** Participants, vaccine effectiveness and long COVID risk reduction during first COVID-19 episode.

| Participants |  |  |  |  |
| --- | --- | --- | --- | --- |
|  | Cases | Controls | Months since vaccination (median (IQR)) | Adjusted VE (%) or risk reduction (%) (95% CI) <sup>a,b</sup> |
| COVID-19 VE analysis |  |  |  |  |
| Case and control definition | COVID-19 cases | Test-negative |  |  |
| NV (pre-Omicron period) <sup>c</sup> | 180 (80.4) | 1254 (56.0) | NA | Reference |
| V1 | 44 (19.6) | 986 (44.0) | 1.6 (0.8 – 2.8) | 75% (64–83) |
| NV (pre-Omicron period) <sup>d</sup> | 44 (36.7) | 299 (24.9) | NA | Reference |
| V2 | 76 (63.3) | 901 (75.1) | 5.4 (3.8 – 6.8) | 95% (84–98) |
| V2_6m (Omicron period) <sup>e</sup> | 702 (14.3) | 1190 (11.3) | 7.9 (7.0 – 9.5) | Reference |
| V3-V5 | 4219 (85.7) | 9376 (88.7) | 2.7 (1.1 – 4.4) | 41% (34–47) |
| Long COVID VE analysis |  |  |  |  |
| Case and control definition | Long COVID cases | Test-negative |  |  |
| NV (pre-Omicron period) <sup>c</sup> | 65 (89.0) | 426 (58.4) | NA | Reference |
| V1 | 8 (11.0) | 304 (41.6) | 1.6 (0.8 – 2.7) | 91% (79–96) |
| NV (pre-Omicron period) <sup>d</sup> | 14 (40.0) | 109 (31.1) | NA | Reference |
| V2 | 21 (60.0) | 241 (68.9) | 5.4 (3.6 – 6.9) | 87% (27–98) |
| V2_6m (Omicron period) <sup>e</sup> | 138 (17.5) | 369 (9.4) | 9.0 (7.5 – 12.5) | Reference |
| V3-V5 | 652 (82.5) | 3559 (90.6) | 3.7 (2.0 – 6.4) | 57% (46–66) |
| Long COVID risk reduction analysis |  |  |  |  |
| Case and control definition | Long COVID cases | COVID-19 controls |  |  |
| NV (pre-Omicron period) <sup>c</sup> | 65 (89.0) | 158 (73.8) | NA | Reference |
| V1 | 8 (11.0) | 56 (26.2) | 2.5 (1.4 – 3.0) | 77% (38–92) |
| NV (pre-Omicron period) <sup>d</sup> | 14 (40.0) | 42 (38.5) | NA | Reference |
| V2 | 21 (60.0) | 67 (61.5) | 5.6 (4.2 – 6.9) | -122% (-2119–78) |
| V2_6m (Omicron period) <sup>e</sup> | 138 (17.5) | 773 (11.9) | 8.0 (6.9 – 10.5) | Reference |
| V3-V5 | 652 (82.5) | 5747 (88.1) | 3.7 (2.0 – 6.2) | 47% (33–57) |

Abbreviations: CI, confidence interval; IQR, interquartile range; NA, non-applicable; NV, not vaccinated; RR, risk reduction; V1-V5, vaccination with one to five doses; V2\_6m, vaccination with two doses ≥6 months before testing date; VE, vaccine effectiveness; W, epi-week.
<sup>a</sup> Logistic regression models conditional to biweekly periods compared vaccinated participants without prior infection versus participants not vaccinated (pre-Omicron period) or vaccinated with two doses 6 months or more before testing (Omicron period), adjusted for age group, sex, region of residence, occupation, workplace, material and social deprivation index, number of comorbidities
<sup>b</sup> Vaccine effectiveness and risk reduction calculated as $(1 - \text{adjusted OR}) \times 100$
<sup>c</sup> W2021-01 to 2021-23
<sup>d</sup> W2021-13 to 2021-49
<sup>e</sup> W2021-51 to 2023-08

During the Omicron period, booster-dose VE (V3-V5 relative to V2) by TND was 41% (95%CI 34–47) against first COVID-19 and 57% (95%CI 46–63) against long COVID post-first infection (Table 2). Pre-Omicron prior infection was associated with hybrid VE against COVID-19 reinfection of 60% (95%CI 51–67); hybrid VE for Omicron prior infection ranged 76% (95%CI 69–81) to 79% (95%CI 75–83) (Figure 2; Supplementary Table 2). Prior Omicron infection was associated with similar hybrid VE against long COVID for two-dose (87% 95%CI 72–94) or booster-dose recipients (85% [95%CI 73–91] to 92% [95%CI 87– 95]) (Figure 2; Supplementary Table 3). For different combinations of prior infection and vaccine doses, risk reduction of long COVID post-reinfection varied between 65% and 79%, with large confidence intervals (Figure 2; Supplementary Table 4).

**Figure 2.**
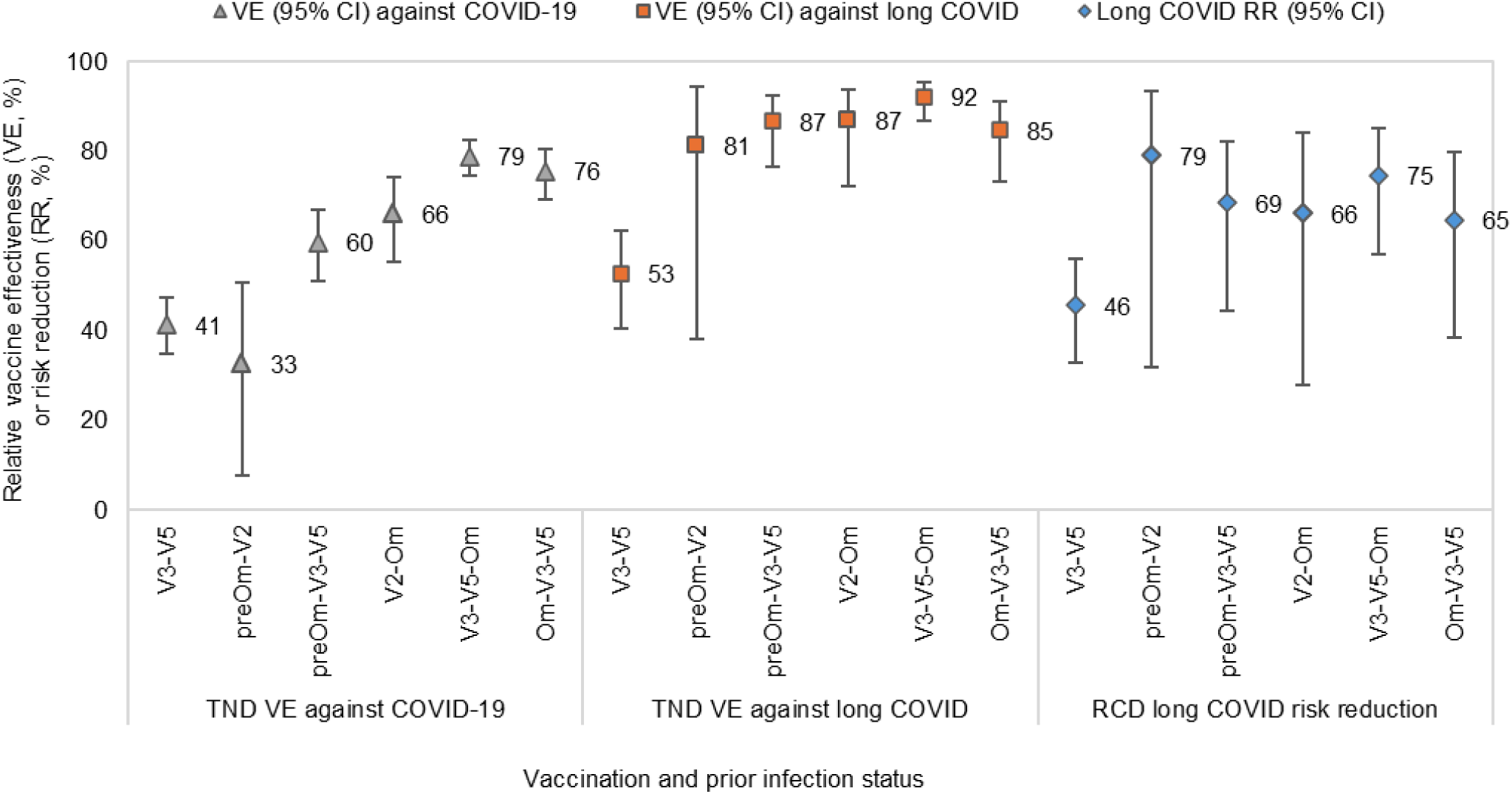
Protection against COVID-19 and long COVID associated with vaccination and prior infection during Omicron period. Logistic regression models compared vaccinated participants with or without prior infection versus participants vaccinated with two doses ≥6 months before testing and without documented or self-reported prior infection. Abbreviations: CI, confidence interval; RCD, retrospective cohort design; RR, risk reduction; TND, test-negative design; V3-V5, vaccination with three to five doses without prior infection; PreOm-V2, prior pre-Omicron infection before two vaccine doses; PreOm-V3-V5, prior pre-Omicron infection before three to five vaccine doses; V2-Om, prior Omicron infection after two vaccine doses; V3-V5-Om, prior Omicron infection after three to five vaccine doses; Om-V3-V5, prior Omicron infection before three to five vaccine doses; VE, vaccine effectiveness.

TND-estimated booster-dose VE against COVID-19 and long COVID among individuals without prior infection steadily decreased over time, becoming low and non-significant at 6 months post-vaccination (13% [95%CI -6–28] against COVID-19 and 29% [95%CI -1–50] against long COVID) (Figure 3). Although the number of long COVID cases post-reinfection was too limited to stratify for time since last immunological event, VE estimates were similar regardless of median time since last vaccination, which ranged from 2 to 9 months) (Supplementary Table 1).

**Figure 3.**
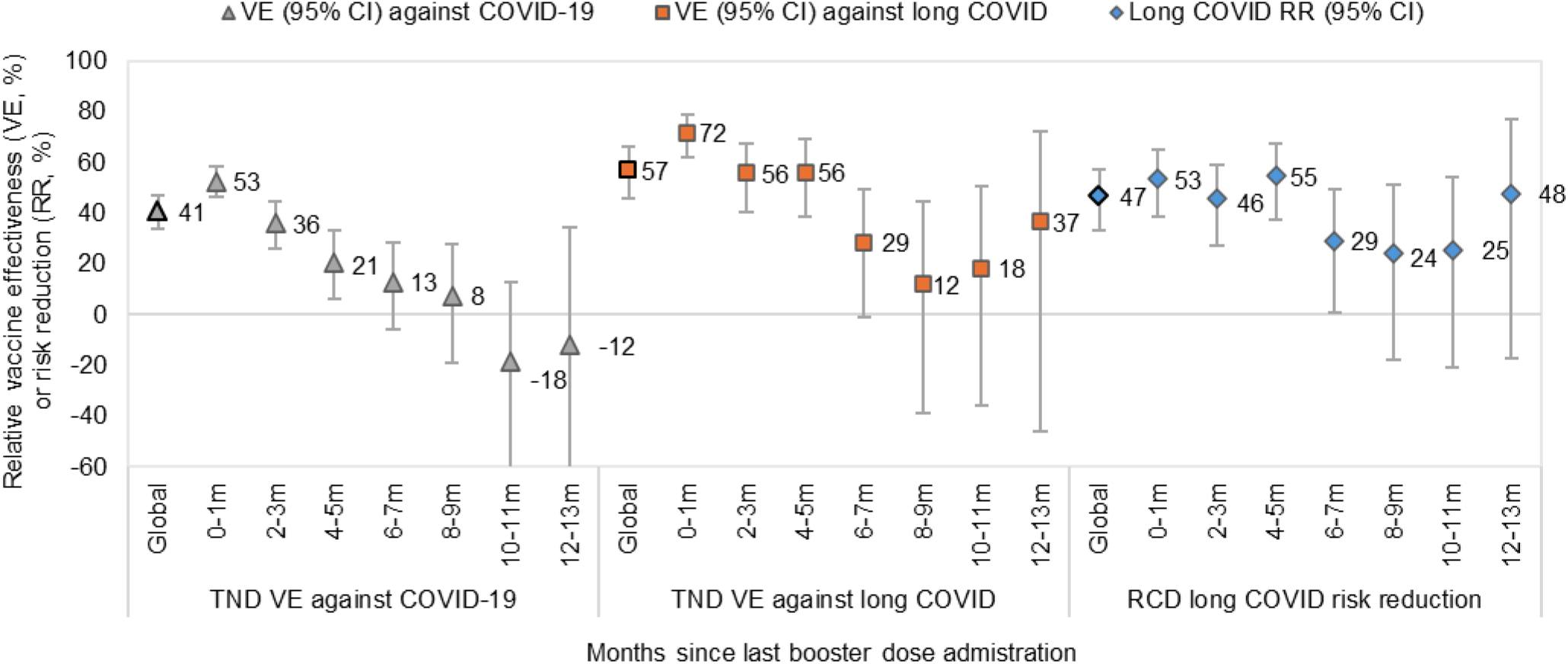
Protection against COVID-19 and long COVID associated with booster vaccination without prior infection, by time since vaccination. Logistic regression models compared participants vaccinated with a booster dose without prior infection versus participants vaccinated with two doses 6 months or more before testing, stratified by time since last booster vaccination. Abbreviations: CI, confidence interval; M, month; RCD, retrospective cohort design; RR, risk reduction; TND, test-negative design; VE, vaccine effectiveness.

In all scenarios, estimated VE against long COVID with moderate or severe symptoms were similar to the protection against long COVID overall (Supplementary Table 5).

## DISCUSSION

In this analysis of vaccine protection against long COVID, we extend earlier findings by revealing the important benefit of preventing COVID-19 infection in the first place. While seemingly obvious to consider in quantifying vaccine benefit against long COVID, most prior analyses have overlooked this critical aspect, considering only the prevention of long COVID among those already infected. During the pre-Omicron period, we show that COVID-19 vaccination reduced the long COVID risk by about 90%, mostly by reducing the risk of COVID-19 disease. During the Omicron period, booster dose VE against long COVID post-first infection was ∼60%, waning at 6 months post-vaccination. Conversely, hybrid VE against long COVID resulting from prior infection and 2-5 vaccine doses was 80 to 90%, and did not vary further based upon the number of doses, prior infecting variant or median time since vaccination, up to 9 months.

Although over 80 studies have examined the impact of vaccination on long COVID [3], most included only SARS-CoV-2-infected participants and reported long COVID risk reductions after breakthrough infections that cannot be interpreted as global VE [4,16,17]. Our study reports vaccine effects against long COVID not only among COVID-19 cases, but also relative to non-cases, capturing the fuller spectrum of vaccine benefit. We also computed VE against COVID-19 per se to contextualize risk reduction estimates. For instance, one-dose VE of 91% against long COVID encompasses also the 75% protection against COVID-19, and among the 25% who had breakthrough COVID-19 infections despite vaccination the additional 77% long COVID risk reduction.

During the pre-Omicron period, two European studies including four countries estimated one-dose effectiveness of 35-58% against long COVID, much lower than our 91% estimate [7,8]. Using administrative health records, they defined long COVID by the presence of ≥1 core long COVID symptoms, but without specific COVID-19 attribution. A previous study using the same data source from UK reported one-dose VE against COVID-19 of 57% to 75% [18]. VE against long COVID was paradoxically lower than against COVID-19, suggesting potential underestimation of VE against long COVID. Studies reporting one-dose long COVID risk reduction among COVID-19 cases did not observe vaccine protection with estimates ranging from -4% to 4% [19–21]. Most studies evaluating the additional protection of a second dose among COVID-19 patients reported long COVID risk reductions of <50%, ranging 22% to 49% [16,19,21–23]. Conversely, we observed 77% reduction in the risk of long COVID associated with one vaccine dose for those with pre-Omicron breakthrough infections, without evidence of additional benefit against long COVID among infected patients vaccinated with two doses. This result has to be interpreted in the context of a delayed administration of the second vaccine dose in Quebec and a very high two-dose VE against COVID-19, leaving few individuals two-dose vaccinated and infected to assess the additional protection against long COVID among COVID-19 cases [24].

During the Omicron period, a Czech nation-wide study based on administrative data and physician-diagnosed long COVID reported low booster VE against long COVID of 20%, contrasted with their estimated VE against severe COVID-19 of 50% to 80%, and declining over the 10-month post-vaccination follow-up [6]. We observed higher VE, at 72% for long COVID and 56% for COVID-19, during the first two months and the 2-4 months post-vaccination, but a similar temporal pattern of waning and loss of protection at 6-8 months. Among Omicron-infected cases, long COVID risk was reduced by half for individuals who received booster doses, but the effect size decreased to 24%-29% by 6-11 months post-vaccination. This is consistent with most published studies reporting long COVID risk reduction of 15%-70% associated with booster doses [25–28], while other studies reported no protection [21,23,29].

The protection conferred by hybrid immunity is now the most relevant given most individuals have already experienced one or more SARS-CoV-2 infections. In our study, hybrid immunity was associated with high effectiveness (>80%) against long COVID during Omicron reinfections, independent of the number of vaccine doses and the likely variant of prior infection. This was higher than estimates from the Czeck study, where hybrid protection exceeded that of vaccination only [6]. In another study, prior infection was also associated with 86% long COVID risk reduction after adjustment for vaccination status and circulating variant [30]. Our study overcomes important limitations of previous studies with its exclusion of pre-vaccination periods [31–34], accurate timing of vaccination relative to infection [35–37], and more specific long COVID definitions (e.g. with post-COVID symptoms attributed to laboratory-confirmed COVID-19) [7]. The inclusion of the pre-vaccination period overestimates VE because variants during the pre-vaccination period are recognized to be associated with higher long COVID risk [30]. Most studies based on administrative data use non-specific definitions, with long COVID rarely diagnosed with ICD-10 codes [38]. Those using specific physician diagnosis may also be confounded by health-seeking behaviour, the latter correlated with the likelihood of both vaccination and long COVID diagnosis [6,39].

Cohort survey-based studies like ours also have limitations. To mitigate misclassification associated with self-reported COVID-19 attributed persistent symptoms, we included only cases prevalent at the time of the survey, with limited recall bias, and laboratory-confirmed COVID-19 episodes and performed a secondary analysis against moderate or severe long COVID, which increases outcome specificity. Low survey participation may be associated with lack of representativeness for the outcome (e.g. severe long COVID cases may be more interested to participate), but protection against moderate/severe long COVID was similar to overall estimates. There were insufficient unvaccinated participants to measure absolute VE during the Omicron period, but based on other studies, individuals last vaccinated ≥6 months earlier were likely unprotected and thus provided a valid comparison group [15,40]. Undocumented prior infections may underestimate VE [41], but HCWs had unrestricted access and were systematically tested during the pre-Omicron period; we also excluded those with self-reported prior infection but lacking NAAT confirmation. Finally, HCWs were only questioned about first occurrence of long COVID. Consequently, only those without long COVID following their first infection were eligible for the assessment of hybrid immunity. Lower host susceptibility to long COVID, instead of vaccine or prior infection induced immunity, may partially explain the observed protection associated with hybrid immunity. Acknowledging these limitations, we also highlight the strengths and major added contribution of our study that combined survey data from more than 14,000 HCWs with registry confirmed vaccination and SARS-CoV-2 infections and evaluated long COVID VE, including hybrid immunity, for more than two years since the start of COVID-19 vaccination not only among COVID-19 cases but taking into account vaccine benefits against long COVID through upfront prevention of COVID-19 per se.

Overall, our findings strongly reinforce that COVID-19 vaccination protects against long COVID, adding to protection against COVID-19 disease. Vaccination plus prior infection provide the highest protection against long COVID post-Omicron infection. Our study confirms the major role of COVID-19 vaccination programs to reduce long COVID risk, mostly during the pre-Omicron period but moderately also since Omicron emergence with ongoing relevance given Omicron descendants continue to circulate. In the current context of highly vaccinated and infected populations, however, our findings of similar hybrid protection against long COVID regardless of the number of booster doses or time since vaccination suggest that repeated administration is unlikely to provide substantial incremental benefit. This observation warrants further investigation and confirmation by others given the important public health, vaccine program, and cost implications.

## Acknowledgments.

The authors thank Stéphanie Grenier et Josiane Rivard (Centre de Recherche du CHU de Québec – Université Laval) for their work at coordinating and implementing the long COVID survey, and Dr Aissatou Fall (Public Health Agency of Canada) for her contributions in the discussion of the study protocol and the preliminary results. Finally, we thank all healthcare workers who generously participated in the survey.

## Author contributions

All the authors had full access to all the data in the study and take responsibility for the integrity of the data and the accuracy of the data analysis. Concept and design: S.C, G.D.S, D. M.S, C.S and D.T. Acquisition, analysis, or interpretation of data: All authors. Drafting of the manuscript: S.C. Critical revision of the manuscript for important intellectual content: All authors. Statistical analyses: J.P., M.O. and S.C. Supervision: G.D.S. and S.C.

## Data availability

The databases used in this study are a property of the “Ministère de la Santé et des Services sociaux du Québec” that was shared with the researchers under the legal mandate of the National Director of Public Health of Quebec under the Public Health Act, precluding data sharing with a third party. Aggregate data are available within the manuscript and the supplementary material.

