## Supplemental material for "Effectiveness of COVID-19 vaccination and prior infections to reduce long COVID risk during the pre-Omicron and Omicron periods"

**Supplementary Appendix**

**Supplementary Table 1. Vaccination and prior infection status of participants included in the analyses of protection during the Omicron period associated with booster doses**

| **COVID-19 VE analysis, TND** | | | |
|---|---|---|---|
| Case and control definition | COVID-19 cases | Test-negative | Months since vaccination (median (IQR)) |
| Total number (W2021-51 to 2023-08) | 4921 | 10566 | |
| V2_6m (no prior infection) | 702 (12.4) | 1211 (9.3) | 7.9 (7.0 – 9.5) |
| V3-V5 (no prior infection) | 4219 (74.6) | 9730 (74.7) | 2.7 (1.1 – 4.4) |
| PreOm-V2 | 69 (1.2) | 153 (1.2) | 8.5 (7.1 – 11.0) |
| PreOm-V3-V5 | 197 (3.5) | 357 (4.8) | 3.0 (1.2 – 5.1) |
| V2-Om | 92 (1.6) | 204 (1.6) | 6.3 (4.1 – 8.9) |
| V3-V5-Om | 230 (4.1) | 756 (5.8) | 5.7 (4.1 – 7.7) |
| Om-V3-V5 | 146 (2.6) | 375 (2.7) | 2.3 (1.3 – 3.6) |
| **Long COVID VE analysis, TND** | | | |
| Case and control definition | Long COVID cases | Test-negative | Months since vaccination (median (IQR)) |
| Total number (W2021-51 to 2023-08) | 851 | 4256 | |
| V2_6m (no prior infection) | 138 (16.2) | 323 (7.6) | 8.7 (7.5 – 12.5) |
| V3-V5 (no prior infection) | 652 (76.6) | 2887 (67.8) | 3.6 (1.9 – 6.2) |
| PreOm-V2 | 3 (0.4) | 42 (1.0) | 7.9 (6.5 – 11.2) |
| PreOm-V3-V5 | 15 (1.8) | 233 (5.5) | 4.0 (2.3 – 6.7) |
| V2-Om | 8 (0.9) | 115 (2.7) | 6.5 (4.5 – 9.2) |
| V3-V5-Om | 18 (2.1) | 455 (10.7) | 5.4 (3.9 – 7.2) |
| Om-V3-V5 | 17 (2.0) | 201 (4.7) | 2.3 (1.4 – 3.6) |
| **Long COVID risk reduction analysis, RCD** | | | |
| Case and control definition | Long COVID cases | COVID-19 controls | Months since vaccination (median (IQR)) |
| Total number (W2021-51 to 2023-08) | 851 | 7376 | |
| V2_6m (no prior infection) | 138 (16.2) | 773 (10.5) | 8.0 (6.9 – 10.5) |
| V3-V5 (no prior infection) | 652 (76.6) | 5747 (77.9) | 3.7 (2.0 – 6.2) |
| PreOm-V2 | 3 (0.4) | 64 (0.9) | 8.8 (6.9 – 11.6) |
| PreOm-V3-V5 | 15 (1.8) | 203 (2.8) | 3.9 (2.3 – 6.5) |
| V2-Om | 8 (0.9) | 104 (1.4) | 8.2 (6.4 – 10.5) |
| V3-V5-Om | 18 (2.1) | 296 (4.0) | 7.4 (5.7 – 9.7) |
| Om-V3-V5 | 17 (2.0) | 189 (2.6) | 3.3 (2.6 – 4.7) |

Abbreviations: IQR, interquartile range; TND, test-negative design; RCD, retrospective cohort design; V3-V5, vaccination with three to five doses without prior infection; PreOm-V2, prior pre-Omicron infection before two vaccine doses; PreOm-V3-V5, prior pre-Omicron infection before three to five vaccine doses; V2-Om, prior Omicron infection after two vaccine doses; V3-V5-Om, prior Omicron infection after three to five vaccine doses; Om-V3-V5, prior Omicron infection before three to five vaccine doses; W, week.

### Supplementary Table 2. Odds ratios of association between vaccination with or without prior infection and COVID-19 disease, estimated by test-negative design

| Exposure | COVID-19 cases | Test-negative | Unadjusted OR[a] (95% CI) | Adjusted OR[a,b] (95% CI) | VE[c] (95% CI) |
|---|---|---|---|---|---|
| Pre-Omicron period (W2021-01 to 2021-23) | | | | | |
| NV | 180 | 1254 | Reference | Reference | Reference |
| V1 | 44 | 986 | 0.22 (0.15–0.32) | 0.25 (0.17–0.36) | 75% (64–83) |
| Pre-Omicron period (W2021-13 to 2021-49) | | | | | |
| NV | 44 | 299 | Reference | Reference | Reference |
| V2 | 76 | 901 | 0.06 (0.02–0.16) | 0.05 (0.02–0.16) | 95% (84–98) |
| Omicron period (W2021-51 to 2023-08) | | | | | |
| V2_6m (no prior infection) | 702 | 1211 | Reference | Reference | Reference |
| V3 (no prior infection) | 3526 | 8716 | 0.70 (0.63–0.77) | 0.61 (0.54–0.67) | 40% (33–46) |
| PreOm-V3 | 159 | 562 | 0.49 (0.40–0.60) | 0.38 (0.31–0.47) | 62% (53–69) |
| V3-Om | 222 | 737 | 0.52 (0.44–0.62) | 0.19 (0.15–0.23) | 81% (77–85) |
| Om-V3 | 29 | 54 | 0.93 (0.58–1.47) | 0.35 (0.22–0.57) | 65% (43–78) |
| V3-V5 (no prior infection) | 4219 | 9730 | 0.75 (0.68–0.83) | 0.59 (0.53–0.65) | 41% (35–47) |
| PreOm-V2 | 69 | 153 | 0.78 (0.58–1.05) | 0.67 (0.49–0.92) | 33% (8–51) |
| PreOm-V3-V5 | 197 | 357 | 0.55 (0.46–0.66) | 0.40 (0.33–0.49) | 60% (51–67) |
| V2-Om | 92 | 204 | 0.78 (0.60–1.01) | 0.34 (0.26–0.45) | 66% (55–74) |
| V3-V5-Om | 230 | 756 | 0.53 (0.44–0.63) | 0.21 (0.17–0.25) | 79% (75–83) |
| Om-V3-V5 | 146 | 375 | 0.71 (0.57–0.87) | 0.24 (0.19–0.31) | 76% (69–81) |

[a]Logistic regression models conditional to biweekly periods compared vaccinated participants with or without prior infection versus participants not vaccinated (pre-Omicron period) or vaccinated with two doses 6 months or more before testing (Omicron period).

[b]Adjusted for age group, sex, region of residence, occupation, workplace, material and social deprivation index, number of comorbidities

[c]Vaccine effectiveness calculated as 1 – adjusted OR

Abbreviations: CI, confidence interval; NV, not vaccinated; V1, one-dose vaccinated; V2, two-dose vaccinated; V3, three-dose vaccinated; V3-V5, vaccination with three to five doses without prior infection; PreOm-V2, prior pre-Omicron infection before two vaccine doses; PreOm-V3, prior pre-Omicron infection before three vaccine doses; PreOm-V3-V5, prior pre-Omicron infection before three to five vaccine doses; V2-Om, prior Omicron infection after two vaccine doses; V3-Om, prior Omicron infection after three vaccine doses; V3-V5-Om, prior Omicron infection after three to five vaccine doses; Om-V3, prior Omicron infection before three vaccine doses; Om-V3-V5, prior Omicron infection before three to five vaccine doses; OR, odds ratio; VE, vaccine effectiveness; W, week;

### Supplementary Table 3. Odds ratios of association between vaccination with or without prior infection and long COVID, estimated by test-negative design

| Exposure | Long COVID cases | Test-negative | Unadjusted OR[a] (95% CI) | Adjusted OR[a,b] (95% CI) | VE[c] (95% CI) |
|---|---|---|---|---|---|
| Pre-Omicron period (W2021-01 to 2021-23) | | | | | |
| NV | 65 | 426 | Reference | Reference | Reference |
| V1 | 8 | 304 | 0.10 (0.04–0.22) | 0.09 (0.04–0.21) | 91% (79–96) |
| Pre-Omicron period (W2021-13 to 2021-49) | | | | | |
| NV | 14 | 109 | Reference | Reference | Reference |
| V2 | 21 | 241 | 0.17 (0.04–0.74) | 0.13 (0.02–0.73) | 87% (22–98) |
| Omicron period (W2021-51 to 2023-08) | | | | | |
| V2_6m (no prior infection) | 138 | 323 | Reference | Reference | Reference |
| V3 (no prior infection) | 527 | 2340 | 0.53 (0.42–0.66) | 0.50 (0.40–0.63) | 50% (37–60) |
| PreOm-V3 | 8 | 198 | 0.10 (0.05–0.20) | 0.09 (0.04–0.19) | 91% (81–96) |
| V3-Om | 18 | 440 | 0.10 (0.06–0.16) | 0.08 (0.05–0.13) | 92% (87–96) |
| Om-V3 | 3 | 36 | 0.20 (0.06–0.64) | 0.15 (0.04–0.49) | 85% (51–96) |
| V3-V5 (no prior infection) | 652 | 2887 | 0.53 (0.43–0.66) | 0.47 (0.38–0.59) | 53% (41–63) |
| PreOm-V2 | 3 | 42 | 0.17 (0.05–0.55) | 0.19 (0.06–0.62) | 81% (38–94) |
| PreOm-V3-V5 | 15 | 233 | 0.15 (0.09–0.26) | 0.13 (0.08–0.24) | 87% (38–94) |
| V2-Om | 8 | 115 | 0.16 (0.08–0.34) | 0.13 (0.06–0.28) | 87% (72–94) |
| V3-V5-Om | 18 | 455 | 0.09 (0.06–0.15) | 0.08 (0.05–0.13) | 92% (87–95) |
| Om-V3-V5 | 17 | 201 | 0.20 (0.12–0.34) | 0.15 (0.09–0.27) | 85% (73–91) |

[a]Logistic regression models conditional to biweekly periods compared vaccinated participants with or without prior infection versus participants not vaccinated (pre-Omicron period) or vaccinated with two doses 6 months or more before testing (Omicron period).

[b]Adjusted for age group, sex, region of residence, occupation, workplace, material and social deprivation index, number of comorbidities

[c]Vaccine effectiveness calculated as 1 – adjusted OR

Abbreviations: CI, confidence interval; NV, not vaccinated; V1, one-dose vaccinated; V2, two-dose vaccinated; V3, three-dose vaccinated; V3-V5, vaccination with three to five doses without prior infection; PreOm-V2, prior pre-Omicron infection before two vaccine doses; PreOm-V3, prior pre-Omicron infection before three vaccine doses; PreOm-V3-V5, prior pre-Omicron infection before three to five vaccine doses; V2-Om, prior Omicron infection after two vaccine doses; V3-Om, prior Omicron infection after three vaccine doses; V3-V5-Om, prior Omicron infection after three to five vaccine doses; Om-V3, prior Omicron infection before three vaccine doses; Om-V3-V5, prior Omicron infection before three to five vaccine doses; OR, odds ratio; VE, vaccine effectiveness; W, week.

### Supplementary Table 4. Odds ratios of association between vaccination with or without prior infection and long COVID disease among COVID-19 participants

| Exposure | Long COVID cases | COVID-19 controls | Unadjusted OR[a] (95% CI) | Adjusted OR[a,b] (95% CI) | Risk reduction[c] (95% CI) |
|---|---|---|---|---|---|
| Pre-Omicron period (W2021-01 to 2021-23) | | | | | |
| NV | 65 | 1254 | Reference | Reference | Reference |
| V1 | 8 | 986 | 0.35 (0.16–0.77) | 0.23 (0.08–0.62) | 77% (38–92) |
| Pre-Omicron period (W2021-13 to 2021-49) | | | | | |
| NV | 14 | 109 | Reference | Reference | Reference |
| V2 | 21 | 241 | 0.94 (0.43–2.05) | 2.22 (0.22–22.19) | -122% (-2119–78) |
| Omicron period (W2021-51 to 2023-08) | | | | | |
| V2_6m (no prior infection) | 138 | 773 | Reference | Reference | Reference |
| V3 (no prior infection) | 527 | 4743 | 0.62 (0.51–0.76) | 0.56 (0.45–0.69) | 44% (31–55) |
| PreOm-V3 | 8 | 165 | 0.27 (0.13–0.57) | 0.22 (0.10–0.46) | 78% (54–90) |
| V3-Om | 18 | 288 | 0.35 (0.21–0.58) | 0.25 (0.15–0.42) | 75% (58–86) |
| Om-V3 | 3 | 44 | 0.38 (0.12–1.25) | 0.25 (0.08–0.84) | 75% (16–92) |
| V3-V5 (no prior infection) | 652 | 5747 | 0.64 (0.52–0.78) | 0.54 (0.44–0.67) | 46% (33–56) |
| PreOm-V2 | 3 | 64 | 0.26 (0.08–0.85) | 0.21 (0.06–0.68) | 79% (32–94) |
| PreOm-V3-V5 | 15 | 203 | 0.41 (0.24–0.72) | 0.31 (0.18–0.55) | 69% (45–82) |
| V2-Om | 8 | 104 | 0.43 (0.21–0.91) | 0.34 (0.16–0.72) | 66% (28–84) |
| V3-V5-Om | 18 | 296 | 0.34 (0.21–0.57) | 0.25 (0.15–0.43) | 75% (57–85) |
| Om-V3-V5 | 17 | 189 | 0.50 (0.30–0.86) | 0.35 (0.20–0.62) | 65% (38–80) |

[a]Logistic regression models compared vaccinated participants with or without prior infection versus participants not vaccinated (pre-Omicron period) or vaccinated with two doses 6 months or more before testing (Omicron period).

[b]Adjusted for age group, sex, region of residence, occupation, workplace, material and social deprivation index, number of comorbidities and four-week periods.

[c]Risk reduction calculated as 1 – adjusted OR

Abbreviations: NV, not vaccinated; V1, one-dose vaccinated; V2, two-dose vaccinated; V3, three-dose vaccinated; V3-V5, vaccination with three to five doses without prior infection; PreOm-V2, prior pre-Omicron infection before two vaccine doses; PreOm-V3, prior pre-Omicron infection before three vaccine doses; PreOm-V3-V5, prior pre-Omicron infection before three to five vaccine doses; V2-Om, prior Omicron infection after two vaccine doses; V3-Om, prior Omicron infection after three vaccine doses; V3-V5-Om, prior Omicron infection after three to five vaccine doses; Om-V3, prior Omicron infection before three vaccine doses; Om-V3-V5, prior Omicron infection before three to five vaccine doses; CI, confidence interval; OR, odds ratio; W, week.

**Supplementary Table 5. Participants, vaccine effectiveness and risk reduction of moderate or severe long COVID associated with vaccination with or without prior infection**

| **Moderate or severe long COVID VE analysis** | | | |
|---|---|---|---|
| Case and control definition | Long COVID cases | Test-negative | VE (%) (95% CI) |
| Pre-Omicron period (W2021-01 to 23 for V1 and W2021-13 to 49 for V2) | | | |
| NV | 54 (90.0) | 354 (59.0) | Reference |
| V1 | 6 (10.0) | 246 (41.0) | 90% (74–96) |
| NV | 13 (40.6) | 104 (32.5) | Reference |
| V2 | 19 (59.4) | 216 (67.5) | 84% (10–97) |
| Omicron period (W2021-51 to 2023-08) | | | |
| V2_6m (no prior infection) | 105 (17.7) | 241 (8.1) | Reference |
| V3-V5 (no prior infection) | 444 (74.7) | 2017 (68.0) | 55% (41–66) |
| PreOm-V2 | 3 (0.5) | 39 (1.3) | 83% (41–95) |
| PreOm-V3-V5 | 12 (2.0) | 145 (4.9) | 83% (67–91) |
| V2-Om | 8 (1.3) | 83 (2.8) | 81% (59–91) |
| V3-V5-Om | 11 (1.9) | 290 (9.8) | 93% (86–96) |
| Om-V3-V5 | 11 (1.9) | 153 (5.2) | 88% (75–94) |
| **Moderate or severe long COVID risk reduction analysis** | | | |
| Case and control definition | Long COVID cases | COVID-19 controls | Adjusted risk reduction (%) (95% CI) |
| Pre-Omicron period (W2021-01 to 23 for V1 and W2021-13 to 49 for V2) | | | |
| NV | 54 (90.0) | 158 (73.8) | Reference |
| V1 | 6 (10.0) | 56 (26.2) | 83% (46–95) |
| NV | 13 (40.6) | 42 (38.5) | Reference |
| V2 | 19 (59.4) | 67 (61.5) | -117% (-2037–78) |
| Omicron period (W2021-51 to 2023-08) | | | |
| V2_6m (no prior infection) | 105 (17.7) | 773 (10.5) | Reference |
| V3-V5 (no prior infection) | 444 (74.7) | 5747 (77.9) | 52% (39–63) |
| PreOm-V2 | 3 (0.5) | 64 (0.9) | 72% (8–91) |
| PreOm-V3-V5 | 12 (2.0) | 203 (2.8) | 66% (37–82) |
| V2-Om | 8 (1.3) | 104 (1.4) | 53% (-2–78) |
| V3-V5-Om | 11 (1.9) | 296 (4.0) | 79% (59–89) |
| Om-V3-V5 | 11 (1.9) | 189 (2.6) | 68% (37–84) |

Logistic regression models compared vaccinated participants with or without prior infection versus participants not vaccinated (pre-Omicron period) or vaccinated with two doses 6 months or more before testing (Omicron period).

Abbreviations: CI, confidence interval; IQR, interquartile range; NV, not vaccinated; V1, one-dose vaccinated; V2, two-dose vaccinated; V3-V5, vaccination with three to five doses without prior infection; PreOm-V2, prior pre-Omicron infection before two vaccine doses; PreOm-V3-V5, prior pre-Omicron infection before three to five vaccine doses; V2-Om, prior Omicron infection after two vaccine doses; V3-V5-Om, prior Omicron infection after three to five vaccine doses; Om-V3-V5, prior Omicron infection before three to five vaccine doses; RR, risk reduction; VE, vaccine effectiveness; W, week.

### Supplementary Figure 1. Methodological summary of the analyses

| 1. Analysis | VE against COVID-19 | VE against long COVID | Vaccine-associated long COVID risk reduction among COVID-19 cases |
|---|---|---|---|
| 2. Study design | Test-negative case-control design | Test-negative case-control design | Retrospective cohort design |
| 3. Exposure | • V1, V2<br>• Booster dose: V3-5<br>• Hybrid immunity: prior infection and V2-V5 | • V1, V2<br>• Booster dose: V3-5<br>• Hybrid immunity: prior infection and V2-V5 | • V1, V2<br>• Booster dose: V3-5<br>• Hybrid immunity: prior infection and V2-V5 |
| 4. Outcome | • COVID-19 disease | • Long COVID<br>• Moderate or severe long COVID | • Long COVID<br>• Moderate or severe long COVID |
| 3. Case and control definition | **COVID-19 cases** (positive tests in symptomatic individuals)<br>**Test-negative controls** (biweekly random selection of negative tests in symptomatic individuals) | **Long COVID cases** (positive test associated with self-reported long COVID)<br>**Test-negative controls** (biweekly random selection of negative tests in symptomatic individuals) | **Long COVID cases** (positive test associated with self-reported long COVID)<br>**COVID-19 controls without long COVID** (positive test not associated with self-reported long COVID) |

Abbreviations: V1-V5, vaccination with one to five doses; VE, vaccine effectiveness.

### Supplementary Figure 2. Population flowchart

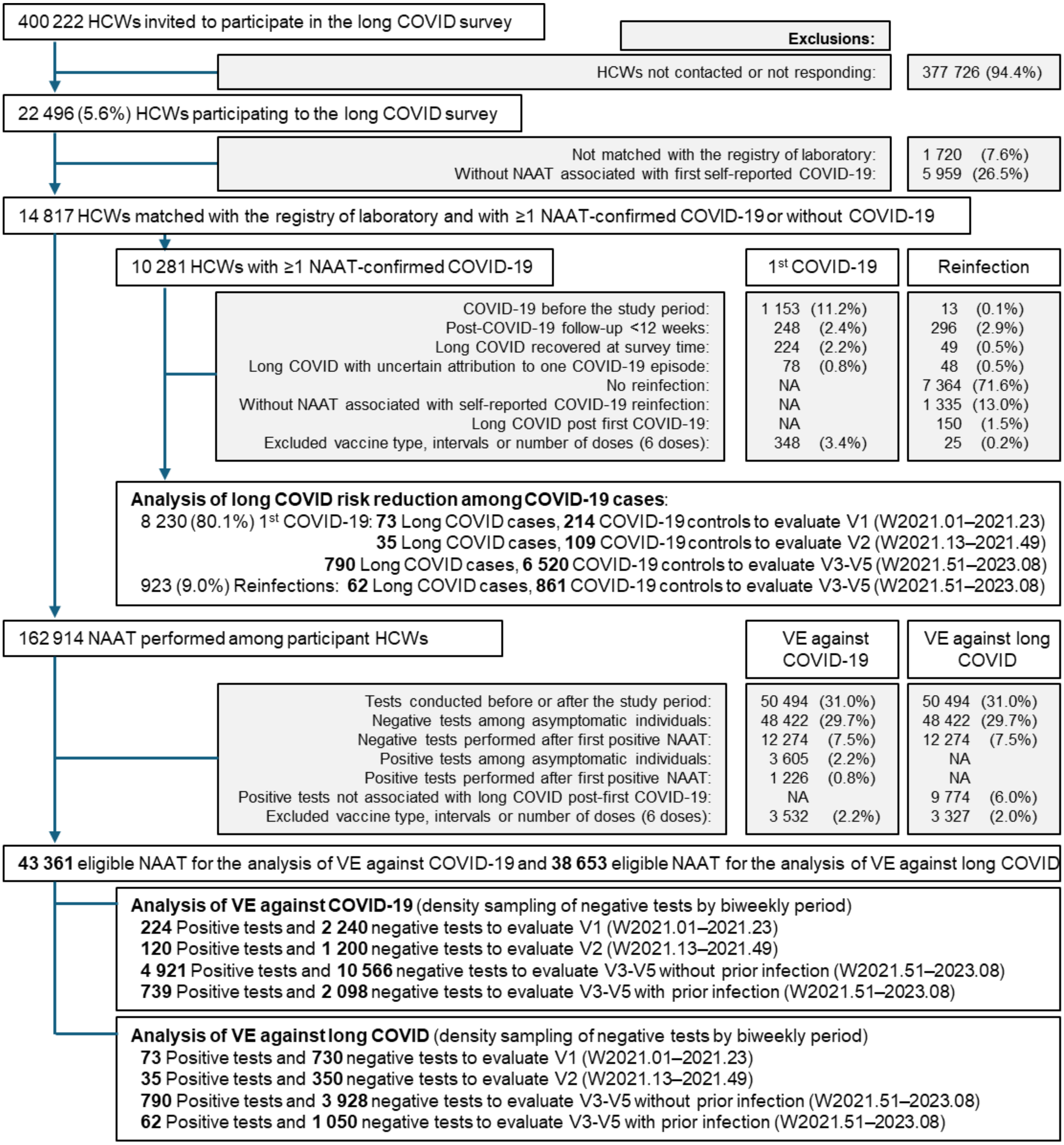

Abbreviations: HCW, healthcare workers. NAAT, nucleic acid amplification test. V1-V5, vaccination with one to five doses. VE, vaccine effectiveness. W, week.
